# United States Life Expectancy Increased in 2023

**DOI:** 10.1101/2024.05.06.24306962

**Authors:** Harry Wetzler

**Affiliations:** Ofstead & Associates, Inc.

## Abstract

From 2019 to 2021 life expectancy at birth in the United States (US) declined by approximately 2.45 years to values not seen since 1996. Although life expectancy increased in 2022, the 2019 levels were not regained. Abridged life tables for 2019 to 2023 were constructed for the total US population, females, and males using mortality data from the CDC WONDER Multiple Cause of Death database and Census Bureau Vintage population estimates. Life expectancy at birth in 2023 increased approximately 1 year compared to 2022. The increase at age 65 was about 0.6 years. The increases in 2022 and 2023 replaced approximately 76% of the pandemic-era declines.

## Introduction

US life expectancies at birth declined by 2.1 years for females and 2.8 years for males between 2019 and 2021.^1,2^ The resultant life expectancies in 2021 were the lowest seen since 1996. Life expectancies at age 65 also declined, 1.1 years for females and 1.2 years for males between 2019 and 2021. My previous results for 2022 and those from CDC/NCHS revealed that approximately 40% of the 2019-2021 declines were regained in 2022.^3^ The purpose of this study was to estimate US life expectancies for 2023.

## Methods

Death data for 2019-2023 by year, gender, and 5-year age groups were obtained from the US Centers for Disease Control and Prevention (CDC) Wide-ranging ONline Data for Epidemiologic Research (WONDER) Multiple Cause of Death database that is updated weekly. This report includes deaths occurring through April 20, 2024 as of April 28, 2024.^4^ Because data were anonymized and publicly available, approval by an institutional review board and informed consent were not required in accordance with 45 CFR §46. Since the 2023 death data are still provisional and incomplete, 4 weeks of data – those for March 30, April 6, 13, and 20, 2024 were modeled to project the number of deaths that will occur in 2023. The equation fitted was:

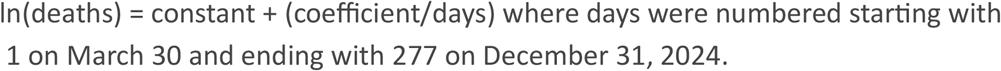

R squared values for all deaths and for females and males separately each exceeded 0.99.

Population data for July 1, 2019 were downloaded from the Census Bureau Vintage 2020 population estimates and data for July 1, 2020-2023 came from the Vintage 2023 estimates.^5,6^ An abridged life table template from Public Health England (PHE) was used for life expectancy and standard error calculations.^7^ The PHE template has 0.12 built in for the fraction of life lived by decedents dying in their first year. This was changed to 0.126, the average of the values used in the 2019-2021 US life tables. An additional nuance of the PHE template is the method for obtaining the years lived in the final open ended age interval for ages 90+. This is calculated by dividing the number surviving by the death rate in the interval. The CDC/NCHS life tables use a blend of vital statistics reported deaths and Medicare data for the oldest ages because Medicare enrollees must have proof of age to enroll and are considered to be more accurate.^2^

Life expectancy estimates obtained from CDC WONDER death and Vintage population data for 2019-2022 were compared to published values from the Centers for Disease Control (CDC) National Center for Health Statistics (NCHS).^1,2,8,9^ The impact of a 0.5% increase in deaths on 2023 life expectancy was calculated. In addition, the number of reduced deaths in 2023 needed to get back to 2019 life expectancy was calculated. Chiang’s formula was used to determine the standard error of life expectancy.^10^ I followed the Strengthening the Reporting of Observational Studies in Epidemiology ([STROBE](http://www.equator-network.org/reporting-guidelines/strobe/)) reporting guidelines.

## Results

As of April 20, 2024, 3,088,797 deaths were reported in the US for 2023, a 5.8% decrease from 2022. The changes for females and males were -5.6% and -6.0% respectively. A total of 3,093,309 deaths were projected to be reported for 2023 by the end of 2024, an increase of 4,512 (0.15%). The standard errors for the 2023 estimates are 0.0085 years at birth and 0.0055 years at age 65. Table 1 lists life expectancies at birth for the published US life tables (CDC/NCHS) and those calculated in this study (the 2022 CDC/NCHS results are reported in tenths of a year).

**Table 1.** Life Expectancies at Birth.

|  |  | Year |  |  |  |  |
| --- | --- | --- | --- | --- | --- | --- |
|  |  | 2019 | 2020 | 2021 | 2022 | 2023 |
| Total | CDC/NCHS | 78.85 | 76.99 | 76.37 | 77.5 | - |
|  | This study | 79.22 | 77.01 | 76.41 | 77.52 | 78.53 |
| Female | CDC/NCHS | 81.39 | 79.88 | 79.33 | 80.2 | - |
|  | This study | 81.78 | 79.81 | 79.36 | 80.24 | 81.16 |
| Male | CDC/NCHS | 76.31 | 74.19 | 73.55 | 74.8 | - |
|  | This study | 76.66 | 74.33 | 73.59 | 74.87 | 75.95 |

In the gender stratified results, the differences between the life expectancies at birth in the CDC/NCHS data and those in this study ranged from -0.40 (−0.49%) years to +0.07 (+0.09%) with a median difference of -0.06 (−0.07%). By far, the largest discrepancies occurred in the year 2019 estimates.

Table 2 lists life expectancies at age 65 for the published US life tables and those calculated in this study.

**Table 2.** Life Expectancies at Age 65.

|  |  | Year |  |  |  |  |
| --- | --- | --- | --- | --- | --- | --- |
|  |  | 2019 | 2020 | 2021 | 2022 | 2023 |
| Total | CDC/NCHS | 19.59 | 18.46 | 18.42 | 18.9 |  |
|  | This study | 20.05 | 18.42 | 18.51 | 19.00 | 19.66 |
| Female | CDC/NCHS | 20.82 | 19.78 | 19.74 | 20.2 |  |
|  | This study | 21.29 | 19.68 | 19.81 | 20.20 | 20.81 |
| Male | CDC/NCHS | 18.20 | 17.03 | 16.99 | 17.5 |  |
|  | This study | 18.65 | 17.06 | 17.09 | 17.67 | 18.37 |

If the final death number for 2023 increased by 0.5% (15,467), then the life expectancy at birth would decrease by 0.06 years, and the decrease at age 65 would be 0.04 years. To reach the pre-pandemic life expectancy at birth in 2019, the 2023 deaths would have had to decrease an additional 5.2%.

The low point in life expectancies at birth occurred in 2021. The subsequent increases replaced approximately 76% of the declines. Life expectancy at age 65 is more complicated. In Table 2 the life expectancy low in the CDC/NCHS data occurred in 2021 whereas it occurred in 2020 for this study’s results. About 77% of the declines were recovered through 2023.

## Discussion

Life expectancies in the US continued to increase in 2023 after starting to rise in 2022. With the gains made in 2022 and 2023, three-fourths of the pandemic era declines have been made up.

Comparing this study’s results with the “official” US results for years 2019-2022 provides a gauge of this study’s accuracy. For 2020-2022, only 2 of the 18 life expectancy differences exceeded 0.1 year. The 2019 differences are greater largely due to the methods used for determining life expectancy in the last interval. Specifically, in 2019 the life expectancies at age 90 in this study exceeded those in the US life tables by an average of 20.3% while the average for 2020 and 2021 was 2.1%. Since the same method was used for each year in this study, it is unclear why the results for 2019 are so different.

The 2023 death data are provisional and incomplete, but Arias and colleagues stated that “Death data are typically more than 99% complete 3 months after the date of death”.^11^ It is noteworthy that the death total jumped by 58,518 deaths (1.93%) between the weeks ending March 30, 2024 and April 6, 2024. However, the increase from April 6 to April 13, 2024, was only 148 deaths and that from April 13 to April 20, 2024, was 303 deaths. The 4,512 added deaths for 2023 result in a decrease in life expectancy at birth of 0.02 years and 0.01 years at age 65.

The 2022 Revision of World Population Prospects prepared by the Population Division of the Department of Economic and Social Affairs of the United Nations Secretariat presents population projections to the year 2100 including life expectancies. Their projection for life expectancy at birth in 2023 in the US, 79.74 years, is considerably greater than the result in this study.^12^

## Conclusion

US life expectancies continued to increase in 2023 although pre-pandemic levels have not been achieved. 75% of the pandemic decline in life expectancy has been made up but it remains to be seen whether the 2019 levels can be achieved in the future.

## Data Availability

All data produced are available online at:
https://wonder.cdc.gov/controller/datarequest/D176
https://www.census.gov/newsroom/press-kits/2020/population-estimates-detailed.html
https://www.census.gov/data/tables/time-series/demo/popest/2020s-national-detail.html

## References

1. Arias E, Xu J. United States Life Tables, 2019. Natl Vital Stat Reports. 2022;70(19). Accessed April 29, 2024. https://www.cdc.gov/nchs/data/nvsr/nvsr70/nvsr70-19.pdf

2. Arias E, Xu J, Kochanek K. United States Life Tables, 2021. Natl Vital Vital Statistics Reports. 2023;72(12). https://www.cdc.gov/nchs/data/nvsr/nvsr72/nvsr72-12.pdf

3. Wetzler H. US Life Expectancy Rebounds in 2022. Published online 2023. 10.1101/2023.02.26.23286363

4. Provisional Mortality Statistics, 2018 through Last Month Request Form. Accessed April 30, 2024. https://wonder.cdc.gov/controller/datarequest/D176

5. US Census Bureau. 2019 Population Estimates by Age, Sex, Race and Hispanic Origin. Accessed April 30, 2024. https://www.census.gov/newsroom/press-kits/2020/population-estimates-detailed.html

6. US Census Bureau. National Population by Characteristics: 2020-2023. Accessed April 30, 2024. https://www.census.gov/data/tables/time-series/demo/popest/2020s-national-detail.html

7. Public Health England. Life Expectancy Template. https://fingertips.phe.org.uk/documents/PHELifeExpectancyCalculator.xlsm

8. Arias E, Xu J. United States Life Tables, 2020. Natl Vital Stat Reports. 2022;71(1). Accessed February 20, 2023. https://www.cdc.gov/nchs/products/index.htm.

9. Kochanek K, Murphy S, Xu J, Arias E. Mortality in the United States, 2022. NCHS Data Brief. 2024;(492). https://www.cdc.gov/nchs/products/databriefs/db492.htm

10. Chiang CL. Life Table and Mortality Analysis. https://apps.who.int/iris/handle/10665/62916

11. Arias E, Kochanek K, Xu J, Tejada-Vera B. Provisional Life Expectancy Estimates for 2022. Natl Cent Heal Stat. 2023;15(31):1–8. https://www.cdc.gov/nchs/data/vsrr/vsrr031.pdf

12. Population Division of the Department of Economic and Social Affairs of the United Nations. World Population Prospects 2022. Published 2022. Accessed May 1, 2024. https://population.un.org/wpp/

